# Association of Oral Triptans and Other Oral Migraine Treatments With Cardiovascular Outcomes in Patients With a History of Cardiovascular Conditions

**DOI:** 10.1101/2025.01.27.25321076

**Authors:** David W. Dodick, Matt Fisher, Maral DerSarkissian, Shawn N. Murphy, Mei Sheng Duh, Chi Gao, Rory B. Weiner, Louise H. Yu, Christopher Herrick, Yichuan Grace Hsieh, Travis Wang, Greg Belsky, Azeem Banatwala, Janet Boyle-Kelly, Jennifer Costello, Marykate Murphy, Amy K. Wong, Jessica Ailani

## Abstract

**OBJECTIVE:** To assess risk of major adverse cardiovascular events associated with acute antimigraine treatments in patients with preexisting cardiovascular conditions.

**PATIENTS AND METHODS:** In this retrospective, longitudinal, observational cohort study, we examined data from the Mass General Brigham Research Patient Data Registry on adults who received one or more prescriptions for acute migraine (index date) between January 2006 and December 2020 and who had one or more diagnoses of a cardiovascular condition during 12 months prior to index date. Study endpoints were nonfatal myocardial infarction, nonfatal stroke, and in-facility all-cause mortality, assessed separately and together as a composite major adverse cardiovascular events–proxy endpoint.

**RESULTS:** Prescriptions were identified for 4016 oral triptans, 6084 opioids/butalbital, and 2021 NSAIDs. Hazard ratios (HRs) for major adverse cardiovascular events were 0.38 (95% CI: 0.22, 0.67; *P*=.001) comparing oral triptans with opioids/butalbital and 0.46 (95% CI: 0.29, 0.71; *P*<.001) with NSAIDs. There was no increased risk of nonfatal stroke comparing oral triptans with opioids/butalbital (HR 0.39; 95% CI 0.22, 0.69; *P*=.001) and with NSAIDs (HR 0.44; 95% CI 0.26, 0.76; *P*=.003). In a subgroup analysis of patients taking sumatriptan, HRs for major adverse cardiovascular events were 0.34 (95% CI: 0.21, 0.56; *P*<.001) when comparing oral sumatriptan with opioids/butalbital and 0.47 (95% CI: 0.28, 0.79; *P*=.004) with NSAIDs.

**CONCLUSIONS:** The risk of adverse cardiovascular events observed with oral triptans is not greater than opioids/butalbital and NSAIDs in patients with migraine and preexisting cardiovascular conditions.

## INTRODUCTION

Migraine headache is widely considered to be associated with trigeminovascular activation and the release of inflammatory and vasomotor neuropeptides, including the calcitonin gene-related peptide (CGRP).^1^ Triptans are selective (5-HT1B/1D) serotonin receptor agonists that have been a standard of care for the acute treatment of moderate or severe migraine headache over the past 3 decades.^2,3^ In addition to lowering levels of CGRP and other neurogenic inflammatory neuropeptides and disrupting trigeminovascular nociceptive transmission,^2,4,5^ triptans may also have off-target effects in the periphery where serotonin receptors are located, such as coronary arteries.^2^ Specifically, by narrowing of epicardial coronary arteries through constriction of smooth muscle and concomitant dilation of coronary resistance vessels through indirect endothelium-dependent nitric oxide-mediated dilation, triptans have the potential to increase the risk of cardiovascular (CV) events.^2,6–9^ Consequently, out of concern for the potential for increased CV risk,^7–9^ initial clinical trials and almost all subsequent pivotal trials of triptans excluded patients with ischemic heart disease, peripheral vascular disease, hypertension, cerebral infarction, or other cardiac impairment.^9–17^ As a result, US prescribing information for triptans identifies contraindications for patients with migraine with a history of CV conditions.^18–20^ Pharmacodynamic studies, however, have found only a modest effect of triptans on coronary arteries *in vitro* and minimal or no effect *in vivo*.^21–23^

In clinical practice, use of nonsteroidal anti-inflammatory drugs (NSAIDs) and opioids is greater than the use of triptans in patients with migraine and CV risk factors,^24^ and butalbital-based therapies remain part of the overall migraine treatment paradigm.^25^ Although triptan use is higher in patients with no CV risk factors than in those with CV risk factors,^26^ as many as 22% of patients prescribed triptans have at least one underlying CV condition considered to be a contraindication for receiving this class of drugs.^27^

The potential risks associated with triptan use compared with other commonly used acute treatments in patients with migraine and a history of CV conditions have not been established.^28^ While several epidemiologic studies examined CV outcomes in patients with migraine who were prescribed triptans, none compared triptans with other therapeutic classes or focused on patients with preexisting CV conditions.^29–32^ A better understanding of the relative risk for CV outcomes associated with oral triptans versus other therapies could help provide insight into their safety profile and inform clinical practice among patients with a history of CV conditions. We hypothesized that the risk of reaching a proxy endpoint for major adverse cardiovascular events (MACE), consisting of nonfatal myocardial infarction (MI), nonfatal stroke, and in-facility all-cause mortality, associated with oral triptans is not different from the risk of reaching a proxy endpoint for MACE for opioids/butalbital-based therapies (O/B) and non-aspirin NSAIDs in patients with migraine and preexisting CV conditions.

## METHODS

### Study design

This was a retrospective, longitudinal, observational cohort study using information extracted from the Mass General Brigham Research Patient Data Registry, which gathers data from eight affiliated hospitals in Massachusetts. The data presented here represent the secondary analysis of previously collected data from a preplanned study of this database. Adult patients prescribed acute antimigraine treatment between January 1, 2006, and December 31, 2020, were identified. The Mass General Brigham Institutional Review Board reviewed (protocol 2021P001844) and allowed an exemption for the study so no informed consent was required. The index date was the date of prescription of antimigraine therapy within 6 months following a migraine diagnosis (**Figure 1**). The baseline period was defined as the 12 months prior to the index date. A washout period of 60 days before the index date was applied, such that antimigraine treatments would not be considered as qualified index prescriptions if there was another antimigraine treatment prescribed in the 60 days prior to the index date. A single patient could contribute data to two separate index dates, provided they were at least 60 days apart, to account for the washout period.

**Figure 1.**
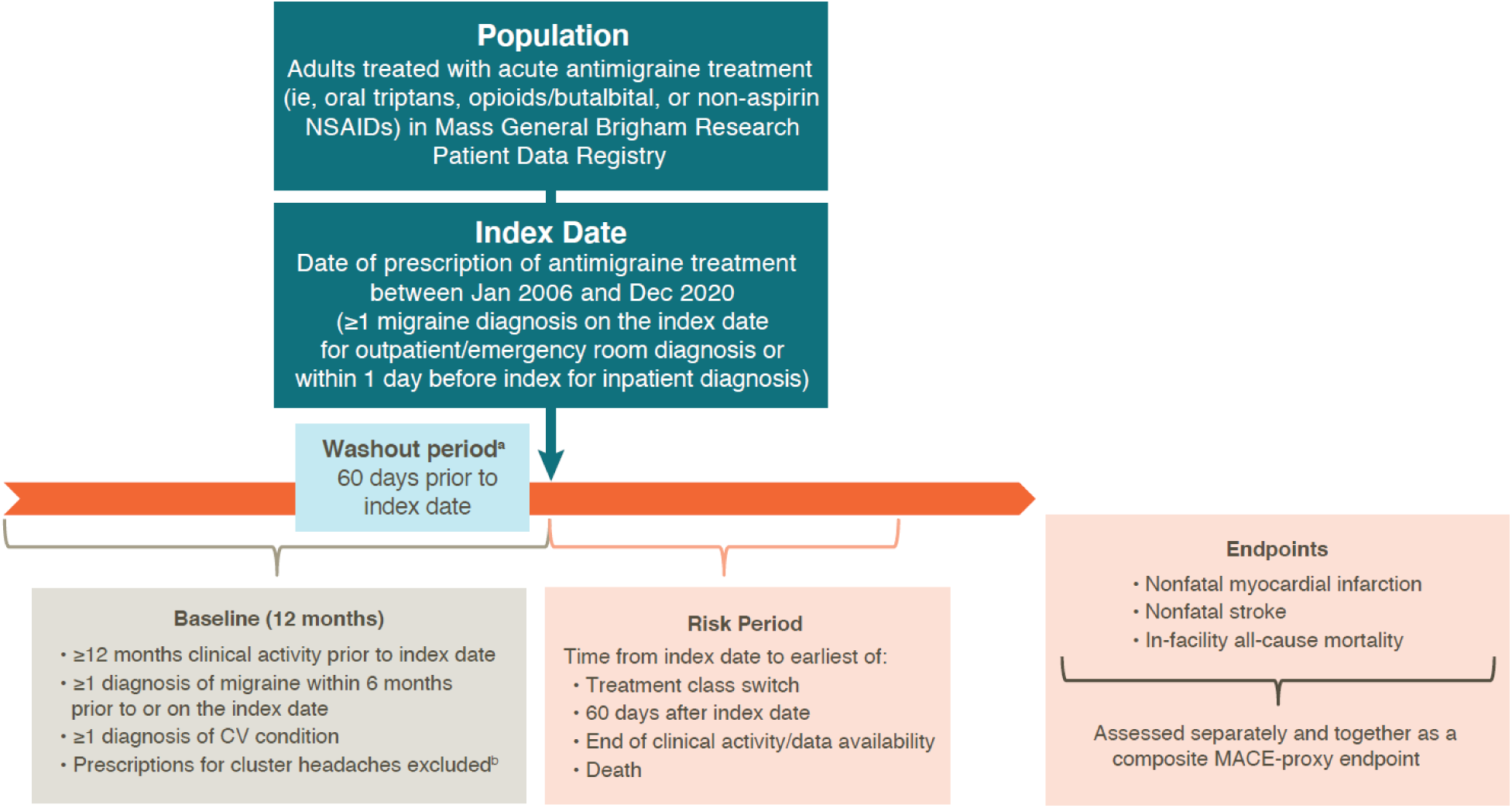
Study design. CV, cardiovascular; MACE, major adverse cardiovascular event; NSAID, nonsteroidal anti-inflammatory drug. ^a^A washout period of 60 days before the index date was applied, such that antimigraine treatments would not be considered as qualified index prescriptions if there was another antimigraine treatment prescribed in the 60 days prior to the index date. ^b^Prescriptions removed if one or more ICD-9-CM/ICD-10-CM codes were associated with a cluster headache diagnosis during baseline or on the index date.

The 12-month baseline period allowed for assessment of patient characteristics and baseline CV risk. The risk period, during which MACE were assessed, was defined as the time from the index date to the earliest of either treatment switching (within or between class), 60 days after the index date, the end of clinical activity, the end of data availability, or death. Patients could contribute more than one risk period if they received more than one eligible prescription.

### Patients

Patients were adults who had received one or more prescriptions for an oral triptan, O/B, or non-aspirin NSAIDs; had two or more outpatient or one or more inpatient migraine diagnoses in any setting within 6 months prior to or on the index date; to ensure that the prescription was written for treatment of migraine, they must have had one or more migraine diagnoses on the index date in the outpatient/emergency room setting or within 1 day of the index date in the inpatient setting; ≥12 months of clinical activity prior to the index date; and one or more ICD-9-CM or ICD-10-CM diagnostic codes associated with a CV condition during the baseline period as described by Dodick et al.^27^ Only oral triptans prescribed as a single antimigraine treatment were included, and combination therapies of O/B and non-aspirin NSAIDs were excluded; combinations of O/B or non-aspirin NSAIDs with other drugs were allowed. Patients with a cluster headache diagnosis during the 12-month baseline period or on the index date were excluded.

### Assessments

The endpoints for this analysis were the occurrence of nonfatal MI, nonfatal stroke, and in-facility all-cause mortality, each assessed separately and together as a composite MACE-proxy endpoint during the risk period (**Figure 1**). In addition, a subgroup analysis was conducted which compared outcomes with oral sumatriptan versus O/B or non-aspirin NSAIDs.

### Statistical analysis

The hazard rate among patients with migraine treated with oral triptans was assumed to be 0.01.^30^ Based on this, it was determined that a minimum total sample of 14,394 evaluable prescriptions was required in the two treatment groups in a 1:1 ratio (7197 per group) to achieve 80% power at a two-sided 5% level of significance to detect a hazard ratio (HR) of 0.8.

Baseline demographic and clinical characteristics were described for the study cohorts using descriptive statistics and are presented as frequencies or mean (standard deviation [SD]) as appropriate. Inverse probability of treatment weighting (IPTW), estimated based on the propensity score (PS), was used to adjust for confounding. The PS was estimated from a logistic regression model that included baseline covariates, including the predicted CV risk score.^33–35^ Variables included in the logistic regression model were covariates that were hypothesized to be associated with both the prescription of antimigraine therapy (ie, the study exposure) and study endpoints and were not mediators of potential effects of interest. The list of variables considered for PS models included demographics (ie, age, sex, race/ethnicity), year of index date, smoking status, body mass index, baseline medication use, Quan-Charlson Comorbidity Index score, CV risk score, baseline CV and non-CV conditions, baseline CV procedures, and number of previous migraine diagnoses. Comorbidities that were not recorded in the patient’s electronic medical records were considered to be absent (ie, the patient did not have the comorbidity). For variables where missing data status could be ascertained (eg, when patient’s body mass index was not reported), we have included an unknown category in the PS models. For variables like comorbidities where absence of information does not indicate unavailable information, only information on presence of comorbidities was used. Patients were not excluded from the study if no comorbidities were observed. Separate PS models were developed for the analysis of oral triptans versus O/B and oral triptans versus non-aspirin NSAIDs. Weights for the oral triptan cohort were calculated as 1/PS, while weights for the reference cohort were calculated as 1/(1−PS) and stabilized by the marginal probability of the respective treatment. The distribution of weights was examined for each comparison (ie, mean [SD] {minimum, maximum}: 0.99 [0.55] {0.45, 16.42} for oral triptans vs O/B-based therapies; 0.99 [0.41] {0.33, 5.46} for oral triptans vs non-aspirin NSAIDs), and no weight trimming was conducted for the main comparative analyses.

After weighting, the distribution of baseline characteristics was evaluated between the cohorts to ensure comparability, and standardized differences were estimated with values >10% indicating an imbalance. Cox proportional hazards models were used to estimate the risk of CV endpoints for oral triptans vs O/B and vs non-aspirin NSAIDs, using an unadjusted and an adjusted model. Results are presented as HRs with 95% CI. Variables that were inadequately balanced between the treatment groups after weighting were added to the IPTW-weighted regression model as covariates. Two-sided tests with a statistical significance level of *P*<.05 were used for hypothesis testing. All analyses were conducted using SAS, version 9.4 (SAS Institute, Cary, NC).

## RESULTS

### Population characteristics

Baseline characteristics of the original (unweighted) population are summarized in **Table 1**. After IPTW, the standardized difference between groups did not exceed 10% for all but one characteristic (**Supplemental Table 1**). A total of 12,121 prescriptions were identified, including 4016 (33.1%) for oral triptans, 6084 (50.2%) for O/B, and 2021 (16.7%) for non-aspirin NSAIDs. The most frequent index treatments were sumatriptan (59.0%) in the oral triptan cohort, butalbital (38.5%), and oxycodone (27.0%) in the O/B cohort, and ibuprofen (61.5%) in the non-aspirin NSAIDs cohort (**Table 2**). The mean ages on the index date were 49.0, 54.2, and 46.6 years for the oral triptan, O/B, and non-aspirin NSAIDs cohorts, respectively; 85.5% to 87.1% of patients in each cohort were female. A greater proportion of African American and Hispanic/Latino patients were prescribed non-aspirin NSAIDs than oral triptans or O/B; correspondingly, fewer White patients were prescribed non-aspirin NSAIDs.

**Table 1.**
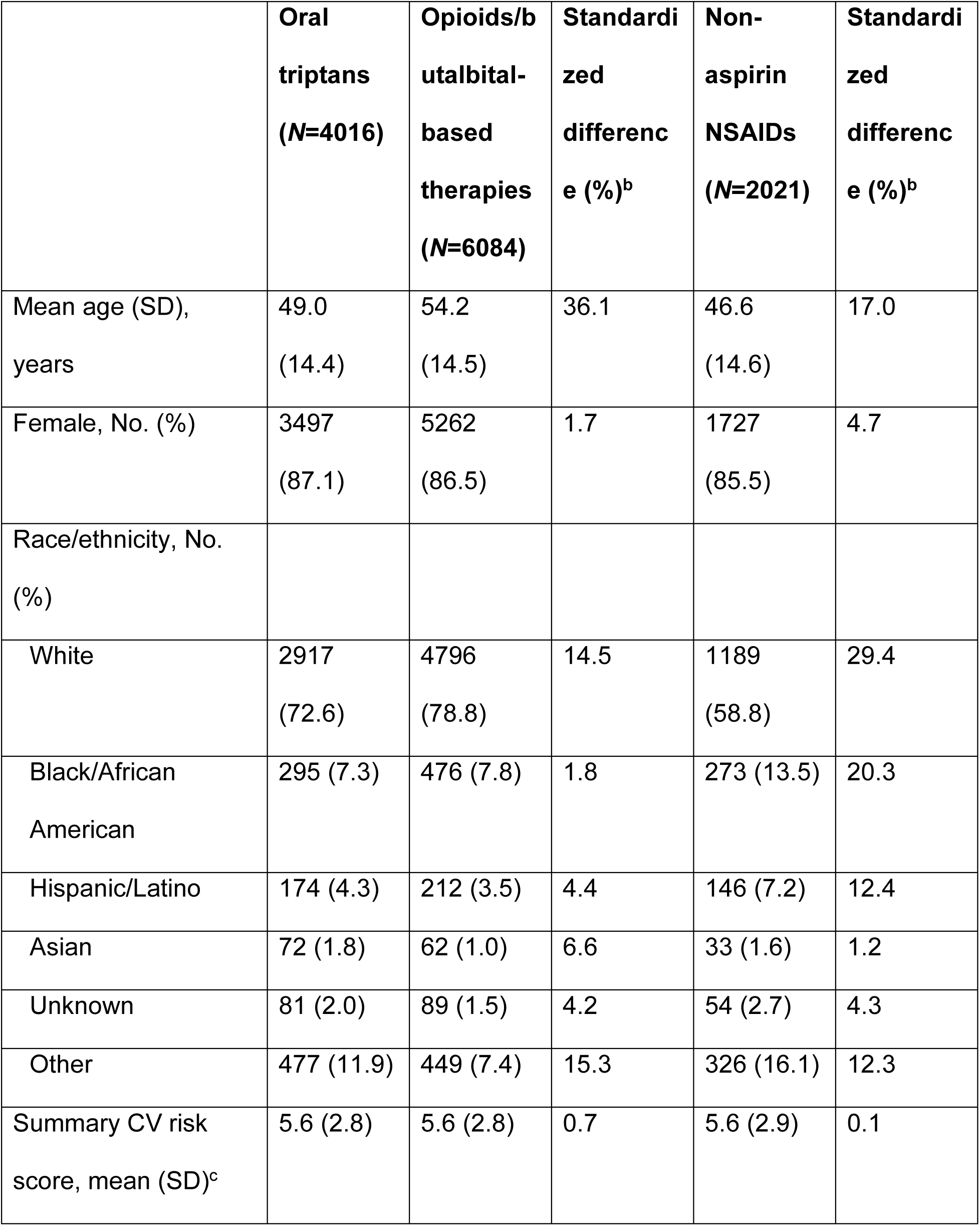
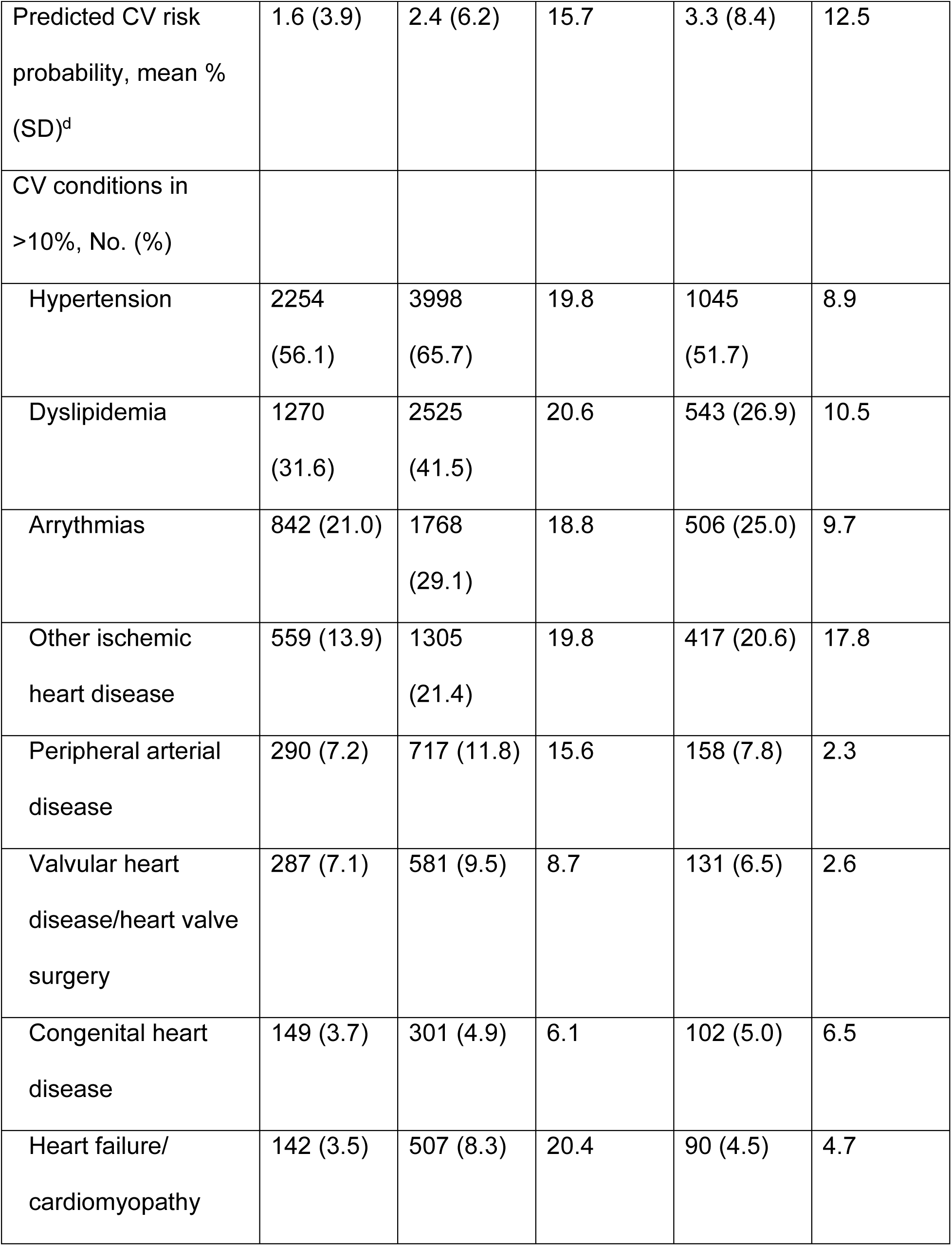
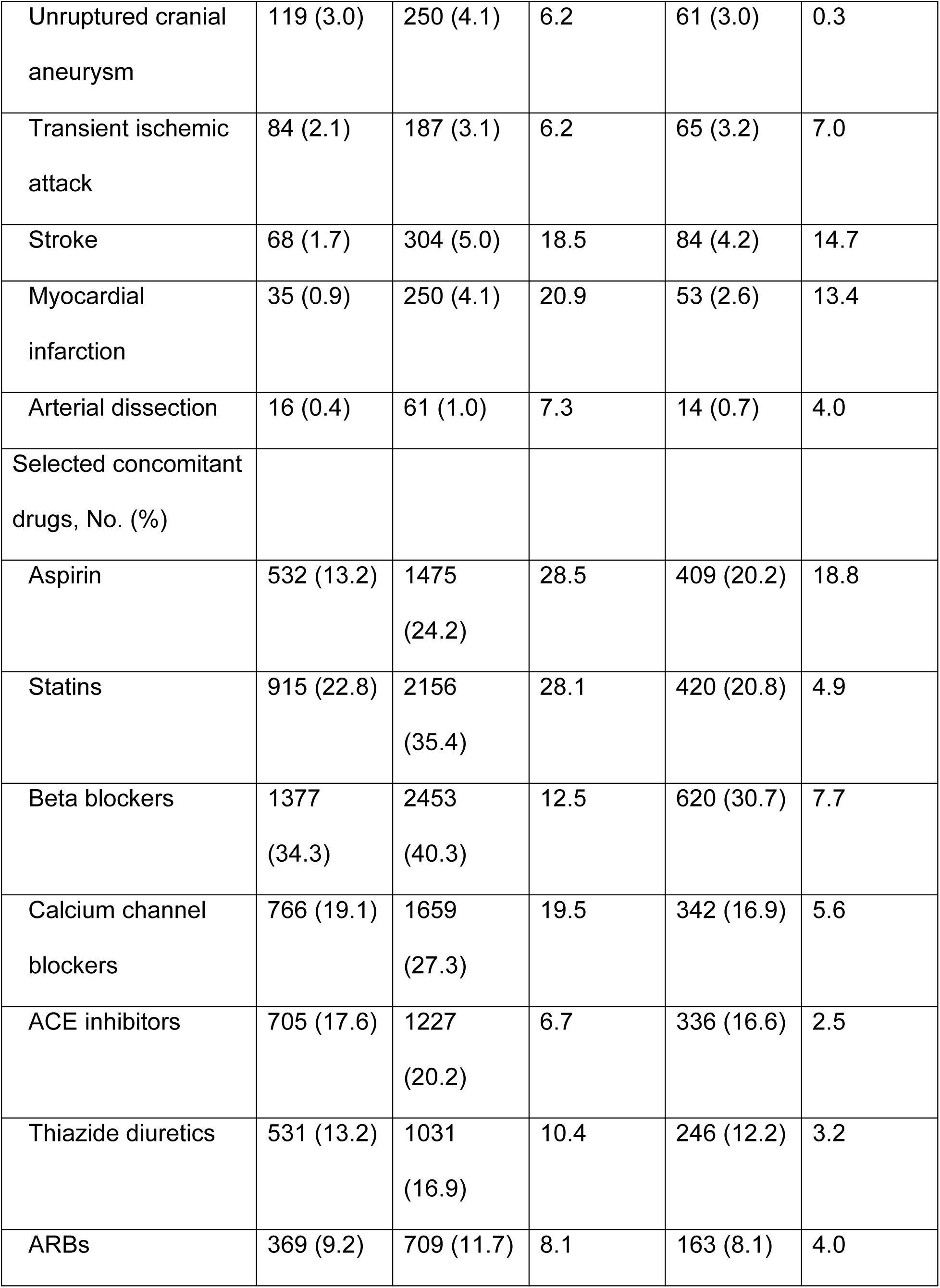
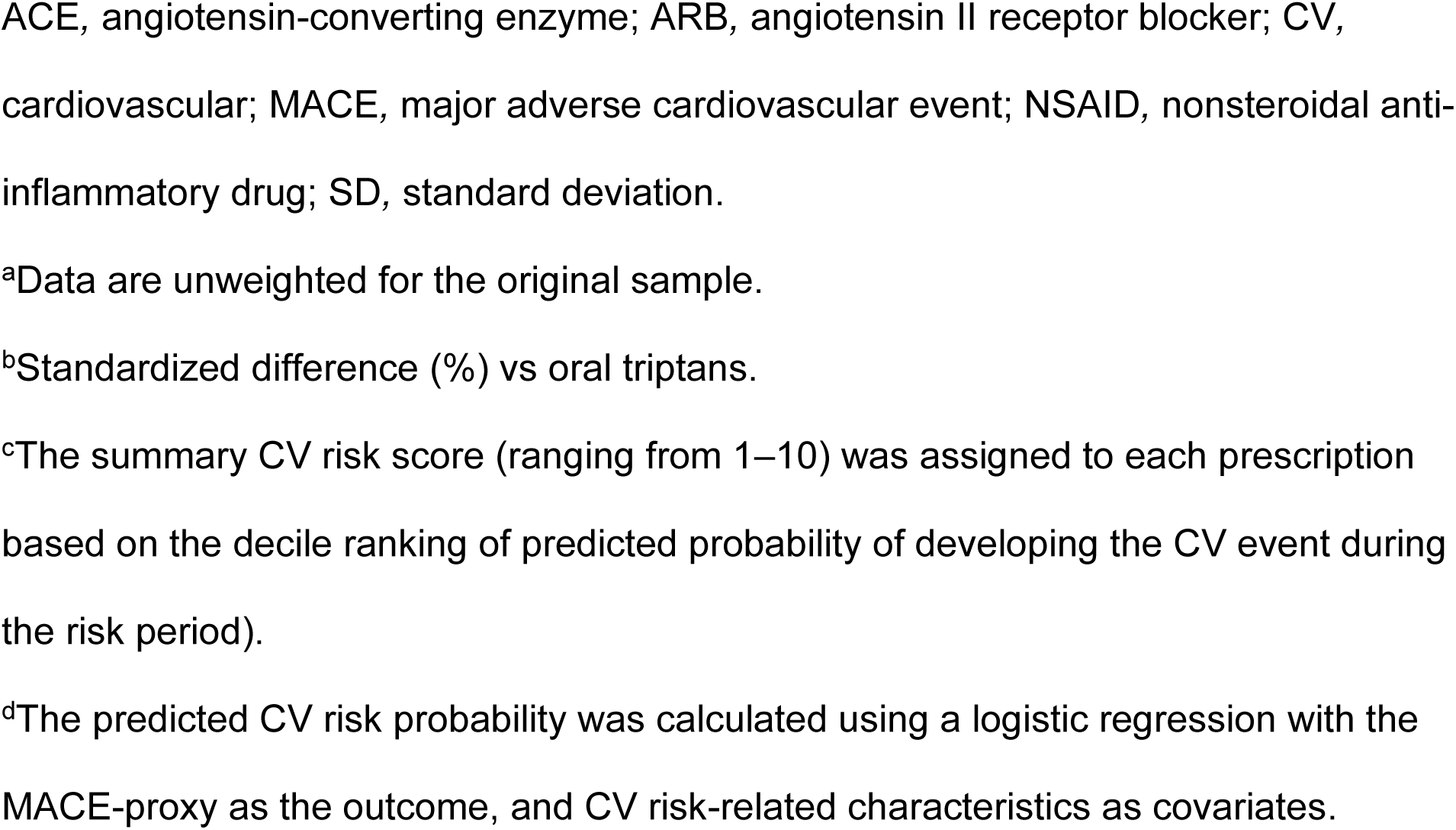
Population characteristics^a^.

|  | **Oral**<br>**triptans**<br>**(N=4016)** | **Opioids/b**<br>**utalbital-**<br>**based**<br>**therapies**<br>**(N=6084)** | **Standardi**<br>**zed**<br>**differenc**<br>**e (%)<sup>b</sup>** | **Non-**<br>**aspirin**<br>**NSAIDs**<br>**(N=2021)** | **Standardi**<br>**zed**<br>**differenc**<br>**e (%)<sup>b</sup>** |
| --- | --- | --- | --- | --- | --- |
| Mean age (SD),<br>years | 49.0<br>(14.4) | 54.2<br>(14.5) | 36.1 | 46.6<br>(14.6) | 17.0 |
| Female, No. (%) | 3497<br>(87.1) | 5262<br>(86.5) | 1.7 | 1727<br>(85.5) | 4.7 |
| Race/ethnicity, No.<br>(%) |  |  |  |  |  |
| White | 2917<br>(72.6) | 4796<br>(78.8) | 14.5 | 1189<br>(58.8) | 29.4 |
| Black/African<br>American | 295 (7.3) | 476 (7.8) | 1.8 | 273 (13.5) | 20.3 |
| Hispanic/Latino | 174 (4.3) | 212 (3.5) | 4.4 | 146 (7.2) | 12.4 |
| Asian | 72 (1.8) | 62 (1.0) | 6.6 | 33 (1.6) | 1.2 |
| Unknown | 81 (2.0) | 89 (1.5) | 4.2 | 54 (2.7) | 4.3 |
| Other | 477 (11.9) | 449 (7.4) | 15.3 | 326 (16.1) | 12.3 |
| Summary CV risk<br>score, mean (SD)<sup>c</sup> | 5.6 (2.8) | 5.6 (2.8) | 0.7 | 5.6 (2.9) | 0.1 |
| Predicted CV risk probability, mean % (SD)<sup>d</sup> | 1.6 (3.9) | 2.4 (6.2) | 15.7 | 3.3 (8.4) | 12.5 |
| CV conditions in >10%, No. (%) |  |  |  |  |  |
| Hypertension | 2254<br>(56.1) | 3998<br>(65.7) | 19.8 | 1045<br>(51.7) | 8.9 |
| Dyslipidemia | 1270<br>(31.6) | 2525<br>(41.5) | 20.6 | 543 (26.9) | 10.5 |
| Arrhythmias | 842 (21.0) | 1768<br>(29.1) | 18.8 | 506 (25.0) | 9.7 |
| Other ischemic heart disease | 559 (13.9) | 1305<br>(21.4) | 19.8 | 417 (20.6) | 17.8 |
| Peripheral arterial disease | 290 (7.2) | 717 (11.8) | 15.6 | 158 (7.8) | 2.3 |
| Valvular heart disease/heart valve surgery | 287 (7.1) | 581 (9.5) | 8.7 | 131 (6.5) | 2.6 |
| Congenital heart disease | 149 (3.7) | 301 (4.9) | 6.1 | 102 (5.0) | 6.5 |
| Heart failure/ cardiomyopathy | 142 (3.5) | 507 (8.3) | 20.4 | 90 (4.5) | 4.7 |
| Unruptured cranial aneurysm | 119 (3.0) | 250 (4.1) | 6.2 | 61 (3.0) | 0.3 |
| Transient ischemic attack | 84 (2.1) | 187 (3.1) | 6.2 | 65 (3.2) | 7.0 |
| Stroke | 68 (1.7) | 304 (5.0) | 18.5 | 84 (4.2) | 14.7 |
| Myocardial infarction | 35 (0.9) | 250 (4.1) | 20.9 | 53 (2.6) | 13.4 |
| Arterial dissection | 16 (0.4) | 61 (1.0) | 7.3 | 14 (0.7) | 4.0 |
| Selected concomitant drugs, No. (%) |  |  |  |  |  |
| Aspirin | 532 (13.2) | 1475 (24.2) | 28.5 | 409 (20.2) | 18.8 |
| Statins | 915 (22.8) | 2156 (35.4) | 28.1 | 420 (20.8) | 4.9 |
| Beta blockers | 1377 (34.3) | 2453 (40.3) | 12.5 | 620 (30.7) | 7.7 |
| Calcium channel blockers | 766 (19.1) | 1659 (27.3) | 19.5 | 342 (16.9) | 5.6 |
| ACE inhibitors | 705 (17.6) | 1227 (20.2) | 6.7 | 336 (16.6) | 2.5 |
| Thiazide diuretics | 531 (13.2) | 1031 (16.9) | 10.4 | 246 (12.2) | 3.2 |
| ARBs | 369 (9.2) | 709 (11.7) | 8.1 | 163 (8.1) | 4.0 |

ACE, angiotensin-converting enzyme; ARB, angiotensin II receptor blocker; CV, cardiovascular; MACE, major adverse cardiovascular event; NSAID, nonsteroidal anti-inflammatory drug; SD, standard deviation.
<sup>a</sup>Data are unweighted for the original sample.
<sup>b</sup>Standardized difference (%) vs oral triptans.
<sup>c</sup>The summary CV risk score (ranging from 1–10) was assigned to each prescription based on the decile ranking of predicted probability of developing the CV event during the risk period).
<sup>d</sup>The predicted CV risk probability was calculated using a logistic regression with the MACE-proxy as the outcome, and CV risk-related characteristics as covariates.

**Table 2.** Prescribed acute antimigraine medications^a^.

| **Oral triptans (N=4016)** | **Opioids/butalbital-based therapies (N=6084)** | **Non-aspirin NSAIDs (N=2021)** |
| --- | --- | --- |
| No. (%) | No. (%) | No. (%) |
| Sumatriptan: 2368 (59.0) | Butalbital: 2342 (38.5) | Ibuprofen: 1243 (61.5) |
| Rizatriptan: 654 (16.3) | Oxycodone: 1642 (27.0) | Naproxen: 361 (17.9) |
| Zolmitriptan: 380 (9.5) | Tramadol: 288 (4.7) | Indomethacin: 138 (6.8) |
| Eletriptan: 241 (6.0) | Hydromorphone: 257 (4.2) | Diclofenac: 92 (4.6) |
| Naratriptan: 221 (5.5) | Hydrocodone: 232 (3.8) | Celecoxib: 74 (3.7) |
| Frovatriptan: 52 (1.3) | Codeine: 218 (3.6) | Ketorolac: 32 (1.6) |
| Almotriptan: 28 (0.7) | Morphine: 75 (1.2) | Other: 40 (2.0) |
| ≥2 triptans: 72 (1.8)<sup>b</sup> | Buprenorphine: 60 (1.0) | ≥2 non-aspirin NSAIDs: 41 (2.0) |
|  | Other: 64 (1.1) |  |
|  | ≥2 opioids/butalbital: 906 (14.9) |  |

NSAID, nonsteroidal anti-inflammatory drug.
<sup>a</sup>Data are unweighted for the original sample.
<sup>b</sup>Two different oral triptans or two different doses of the same triptan on the index date were allowed.

Compared with the oral triptan cohort (13.2%), the O/B and non-aspirin NSAIDs cohorts had higher baseline usage of low-dose aspirin (24.2% and 20.2%, respectively) and the O/B cohort had higher baseline usage of antihyperlipidemics, including statins (37.9%) compared with the non-aspirin NSAIDs and oral triptan cohorts (22.4% and 24.4%, respectively). Similarly, patients in the O/B cohort had a higher baseline usage of antihypertensive agents (**Table 1**). The predicted CV risk was lower in the oral triptan cohort (1.6%) than in either of the O/B or NSAIDs cohorts (2.4% and 3.3%, respectively). In addition, there were higher rates of hypertension, dyslipidemia, and other CV conditions in the O/B cohort than in the triptan or NSAIDs cohorts (**Table 1**). The average duration of the risk period was 49 days for the total population (52 days for oral triptans, 46 days for O/B, and 50 days for non-aspirin NSAIDs).

Sumatriptan was the most prescribed treatment for acute migraine in this population. Of the 4016 oral triptan prescriptions, 2368 sumatriptan prescriptions (59.0%) were identified (**Table 2**). Baseline characteristics for this subgroup population were consistent with the overall oral triptan population, including CV risk, comorbid conditions, and concomitant medications.

### Clinical endpoints

Oral triptans were associated with fewer MACE-proxy outcomes, in-facility all-cause mortality, nonfatal stroke, and nonfatal MI, compared with O/B and non-aspirin NSAIDs (**Figure 2**). The unadjusted HRs for all comparisons of oral triptan vs O/B or non-aspirin NSAIDs were statistically significant (*P*<.001), except all-cause mortality which could not be assessed due to an absence of events in the oral triptan cohort.

**Figure 2.**
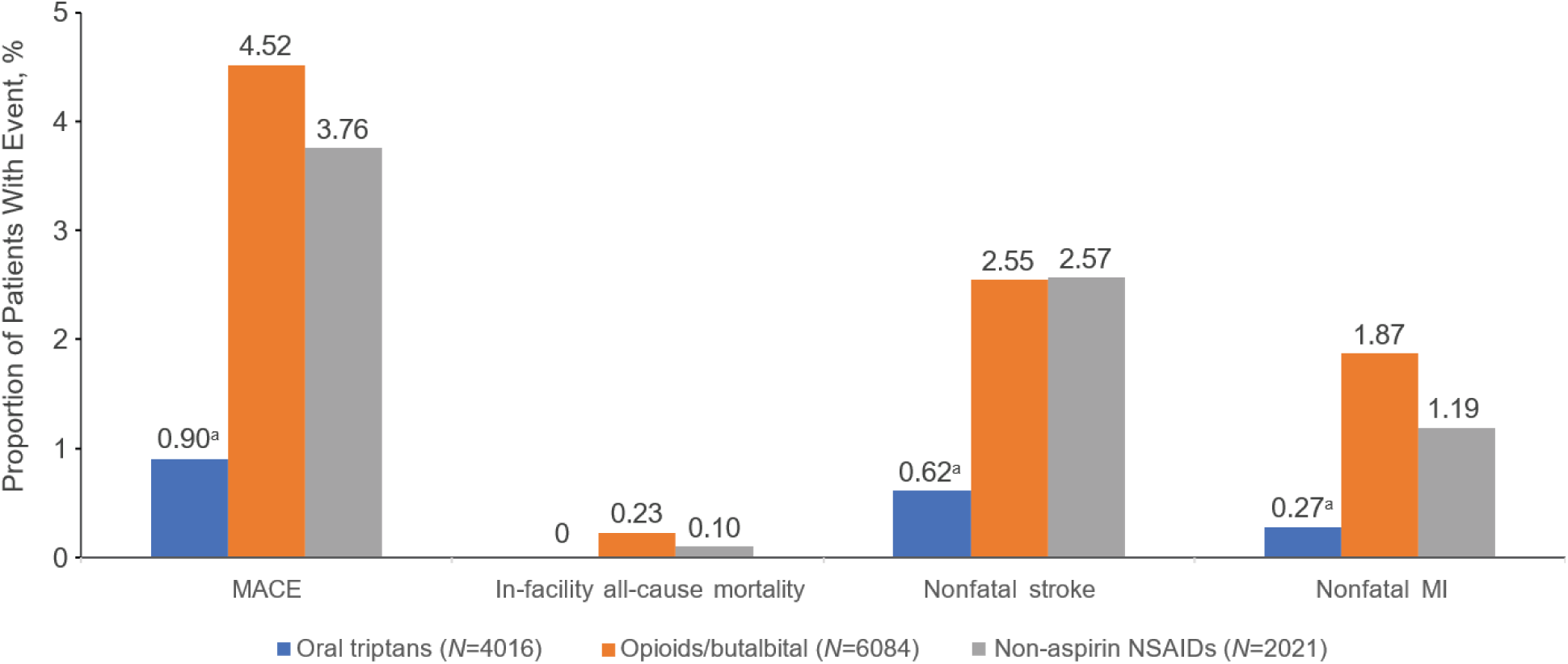
Summary of cardiovascular endpoints during the risk period (unweighted estimates). MACE, major adverse cardiovascular event (defined as first occurrence of any nonfatal MI, nonfatal stroke, or in-facility all-cause mortality); MI, myocardial infarction; NSAID, nonsteroidal anti-inflammatory drug. ^a^*P*<.001 vs opioids/butalbital-based therapies and non-aspirin NSAIDs.

In the adjusted model, HRs for MACE-proxy outcomes were 0.38 (95% confidence interval [CI]: 0.22, 0.67) when comparing oral triptans with O/B and 0.46 (95% CI: 0.29, 0.71) when comparing oral triptans with non-aspirin NSAIDs (*P*≤.001; **Figure 3**). There was also no increased risk of nonfatal stroke with oral triptans vs O/B vs non-aspirin NSAIDs (*P*≤.003). The risk of nonfatal MI was lower with oral triptans compared to O/B and non-aspirin NSAIDs, but this did not reach statistical significance (**Figure 3**). The absence of any in-facility all-cause mortality events in the oral triptan cohort meant that statistical comparisons were not possible.

**Figure 3.**
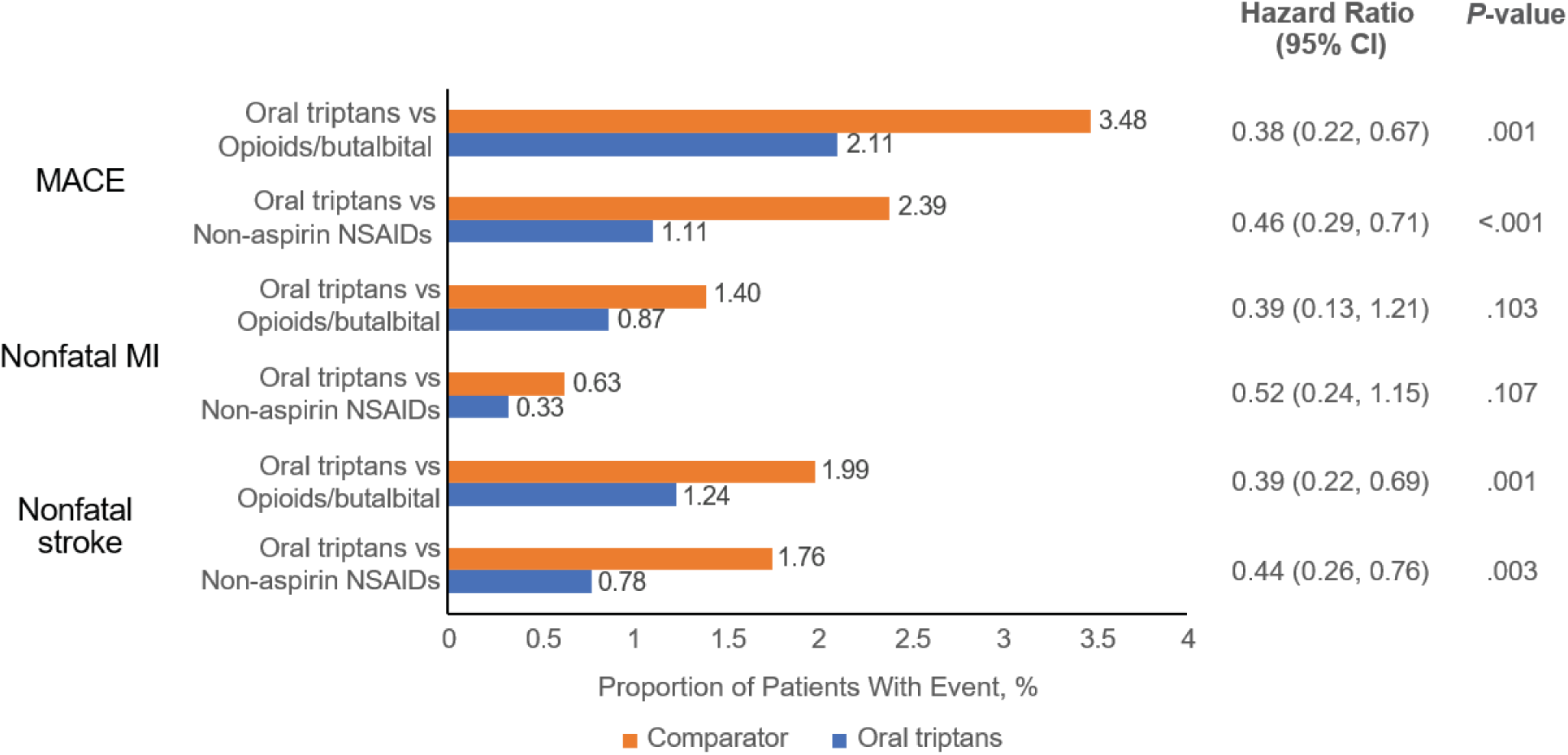
Proportion of patients with cardiovascular events during the risk period (weighted estimates): oral triptans vs opioids/butalbital and non-aspirin NSAIDs (adjusted model). MACE, major adverse cardiovascular event (defined as first occurrence of any nonfatal MI, nonfatal stroke, or in-facility all-cause mortality); MI, myocardial infarction; NSAID, nonsteroidal anti-inflammatory drug.

### Sumatriptan subgroup analysis

MACE-proxy endpoints occurred in 1.1%, 4.5%, and 3.8% of oral sumatriptan, O/B, and non-aspirin NSAIDs prescriptions, respectively. Overall, there was no increased risk of CV events with oral sumatriptan vs O/B and non-aspirin NSAIDs (*P*≤.015, all comparisons, except all-cause mortality due to absence of events in the oral sumatriptan cohort) (**Supplemental Figure 1**). In the adjusted model, the HRs for MACE-proxy endpoints were 0.34 (95% CI: 0.21, 0.56; *P*<.001) for oral sumatriptan vs O/B and 0.47 (95% CI: 0.28, 0.79; *P*=.004) for oral sumatriptan vs non-aspirin NSAIDs; similar results were obtained for MI, nonfatal stroke, and in-facility all-cause mortality, although the comparison did not reach statistical significance for nonfatal MI with oral sumatriptan vs non-aspirin NSAIDs (**Supplemental Figure 2**).

## DISCUSSION

This large, retrospective, longitudinal cohort study assessed the risk of MACE associated with oral triptans compared with other classes of drugs commonly used for the acute treatment of migraine headache in patients with preexisting CV conditions such as heart failure, valvular heart disease, MI, stroke, and peripheral artery disease. These real-world data show that, despite contraindications against their use in patients with preexisting CV conditions, oral triptans are being used in clinical practice in patients with migraine and a history of CV conditions and are not associated with an increased risk of MACE-proxy endpoints compared with O/B and non-aspirin NSAIDs. A significant reduction was observed for MACE-proxy endpoints with oral triptans compared with O/B or non-aspirin NSAIDs. In addition, there was no evidence of increased risk of nonfatal stroke associated with oral triptans compared with O/B or non-aspirin NSAIDs; a similar result was seen for nonfatal MI, but this did not reach statistical significance in the adjusted model. A subgroup analysis including only oral sumatriptan showed similar results. These data are consistent with a study that found no difference in CV comorbidity between triptan users and nonusers over 50 years of age, suggesting that triptan use does not increase vascular risk in that population.^36^

The contraindication against the use of triptans in patients with CV conditions is largely based on their ability to bind to serotonin (5HT_1B_) receptors on vascular beds.^2,6–9^ However, extensive clinical experience has demonstrated a relatively small number of cases reporting CV events in patients taking sumatriptan during the past 30 years since its initial approval in 1992.^18,37–50^ Several studies that assessed the CV risk associated with triptans employed heterogeneous populations, included patients with preexisting CV conditions, and found little to no risk of CV events, including MI, heart failure, stroke, serious ventricular arrythmia, unstable angina, transient ischemic attack, and death.^29–32^ The results of the present study, therefore, are consistent with these earlier studies.

The apparent lack of increased risk of CV events with oral triptans observed in this study is consistent with a number of studies designed to investigate their biological effects on blood vessels. One study demonstrated no changes in contractility with triptans in patients with and without coronary artery disease.^21^ Indeed, it has been suggested that the reactivity of coronary arteries to sumatriptan is impaired in patients with endothelial dysfunction.^51^ Another study showed that plasma concentrations of eletriptan—equivalent to three to five times that achieved with a supratherapeutic 80-mg oral dose—caused only mild, clinically insignificant reductions in coronary artery diameter, and was not different when compared to placebo.^22^ Consistent with this observation, positron emission tomography studies showed that sumatriptan had no significant effect on myocardial perfusion in female patients with migraine and at a low risk of coronary artery disease.^23^ A study of frovatriptan in patients with migraine with, or at high risk of, coronary artery disease found no effect of the drug on the rates of electrocardiogram changes, ischemia, or arrythmia.^52^

The data presented here should be considered in the context of available treatment options for acute migraine relief, and the risks and benefits of oral triptan use in patients with preexisting CV conditions should be weighed in the context of these new data. The risks associated with other antimigraine treatment classes, including the potential for opioid abuse and overdose and gastrointestinal and renal effects associated with NSAIDs, should also be considered.^53,54^ While non-aspirin NSAIDs are recommended for treatment of mild to moderate migraine, triptans are preferred for moderate to severe migraine.^5^ Given the lower rate of CV events observed in this study that were associated with oral triptans compared with non-aspirin NSAIDs and O/B-based therapies, consideration should be given to preferential use of oral triptans or newer acute treatment options, including CGRP receptor antagonists or serotonin selective 1F receptor agonists in adults with preexisting CV conditions if treatment options are limited. Furthermore, treatment guidelines no longer recommend use of O/B for management of acute migraine; replacing O/B with oral triptans or newer acute treatment options not associated with vasoconstriction nor contraindicated in persons with CV risk should be considered.^5,55–57^

This was a large observational study based on the Mass General Brigham Research Patient Data Registry that adopted a rigorous study design, stringent patient selection criteria, and robust statistical methods. There were some disparities between the oral triptan, O/B, and non-aspirin NSAIDs groups in terms of race, baseline CV risk, and baseline medication use, including antihyperlipidemics and antihypertensives.

These differences may have been due to channeling bias in which patients with lower CV risk were preferentially prescribed oral triptans, and vice versa.^58^ However, the study design and statistical methodology was designed to minimize any such effects. First, it should be noted that all patients included in this study had documented CV conditions at baseline. Second, IPTW was employed to adjust for observed confounders, and the distribution of baseline CV conditions was <10% of the standardized differences between cohorts postadjustment. Finally, the study included patients taking non-aspirin NSAIDs rather than all NSAIDs to avoid the potential impact of aspirin on CV risk and to ensure that the comparator populations were as similar as possible.

A limitation of the current study, and similar studies, is that medication prescription data do not provide definitive information on drug dispensing, adherence, or compliance, which may lead to exposure misclassification. In addition, over-the-counter NSAIDs use was not captured since only prescription information was available in the research patient data registry, potentially also resulting in exposure misclassification.

Although IPTW was used to balance study cohorts on observed baseline characteristics, there may exist residual or unmeasured confounding. Certain variables that influence the choice of treatment and endpoints of interest, such as migraine severity, were not captured in the database. In addition, although a population with preexisting CV disease was selected, it was not possible to adjust for severity of the underlying CV disease. Therefore, there may be residual selection bias such that the oral triptan group may consist of patients with less severe CV disease, though this cannot be ascertained using available data. Other limitations are that reliance on diagnosis and procedure codes to measure outcomes may lead to misclassification or underreporting of patients with migraine, covariates, and CV endpoints, and the 60-day risk period may not inform long-term risks of the assessed antimigraine treatments. For instance, given the nature of electronic medical records, there may be underascertainment of comorbidities if the patient received care outside of the Mass General Brigham system prior to receiving antimigraine treatment within Mass General Brigham. Since this observational study covers a period from 2006 to 2020, it does not include newer classes of antimigraine treatments, including the serotonin 5-HT1F receptor agonist lasmiditan and the CGRP receptor antagonists. These newer agents are not associated with vasoconstriction in preclinical models or contraindications in patients with CV disease,^57,59^ although the long-term CV effects of these agents across a broad range of patient populations have yet to be clearly delineated. Finally, while internal validity of the registry data is high (eg, all programs for data analyses were reviewed and audited by a second analyst to ensure coding accuracy), the patients in the registry used in this study are limited to Massachusetts and may not be demographically representative of the state (eg, patients in the Mass General Brigham system are more likely to be white and have higher socioeconomic status compared with the rest of the Massachusetts patient population) or the United States. Therefore, the results may not be generalizable to all US patients.

## CONCLUSIONS

In this analysis of real-world data, oral triptans were not associated with an increased risk of MACE overall, nonfatal MI, and nonfatal stroke in patients with migraine and a history of CV conditions compared with other antimigraine treatments. A subgroup analysis including only oral sumatriptan showed similar results to those of the oral triptan cohort compared with other antimigraine treatments. These results suggest consideration may be needed when weighing the benefits and risks of antimigraine treatments for adults with preexisting CV conditions. Decreasing barriers to effective acute antimigraine therapy by reducing possible misperceptions regarding the CV safety of oral triptans will help reduce the burden of migraine, which remains an undertreated and debilitating disease.

## DECLARATIONS

### Ethics approval and consent to participate

The Mass General Brigham Institutional Review Board reviewed (protocol 2021P001844) and allowed an exemption for the study so no informed consent was required.

### Availability of data and materials

Anonymized individual participant data and study documents can be requested for further research from https://www.gsk-studyregister.com.

### Authors’ contributions

Study design: MF, MD, MSD, CG, TW, LHY

Participated in the study: NA

Acquisition, analysis, or interpretation of data: All authors

Drafting of the manuscript: All authors

Critical review and revision of the manuscript: All authors

Statistical analysis: MD, MSD, CG, TW, LHY

Final approval of manuscript: All authors

All authors had full access to the data and take full responsibility for the integrity of the data and the accuracy of the data analysis.

## Abbreviations

ACE: angiotensin-converting enzyme
ARB: angiotensin II receptor blocker
CGRP: calcitonin gene-related peptide
CI: confidence interval
CV: cardiovascular
HR: hazard ratio
IPTW: inverse probability of treatment weighting
NSAIDs: nonsteroidal anti-inflammatory drug
MACE: major adverse cardiovascular events
MI: myocardial infarction
O/B: opioids/butalbital-based therapies
PS: propensity score
SD: standard deviation

## Acknowledgments

Medical writing and editorial assistance were provided by James Street, PhD, of Peloton Advantage, LLC, an OPEN Health company, and funded by Haleon (formerly GSK Consumer Healthcare).

## FIGURE LEGENDS

**Supplemental Table 1.**
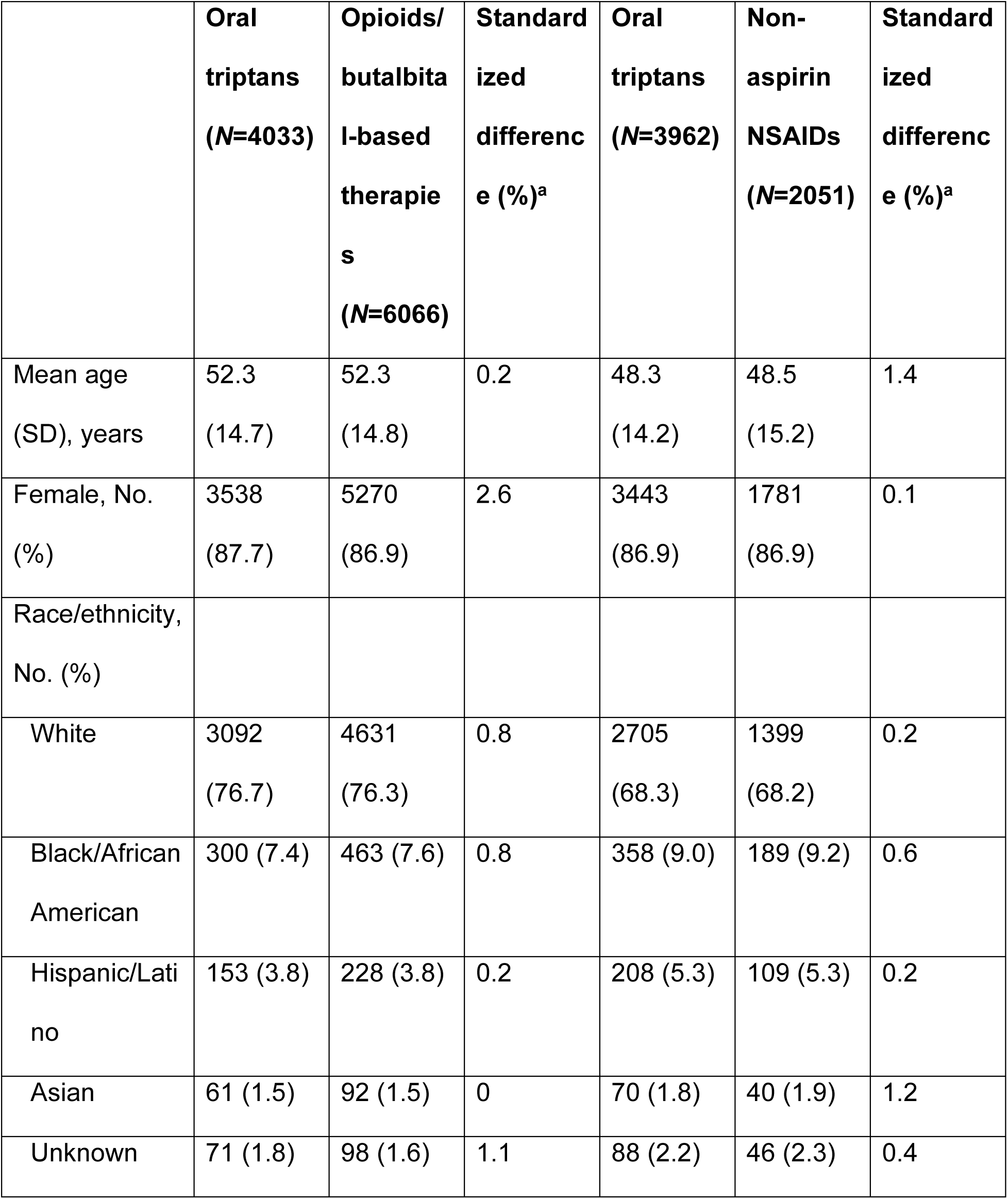
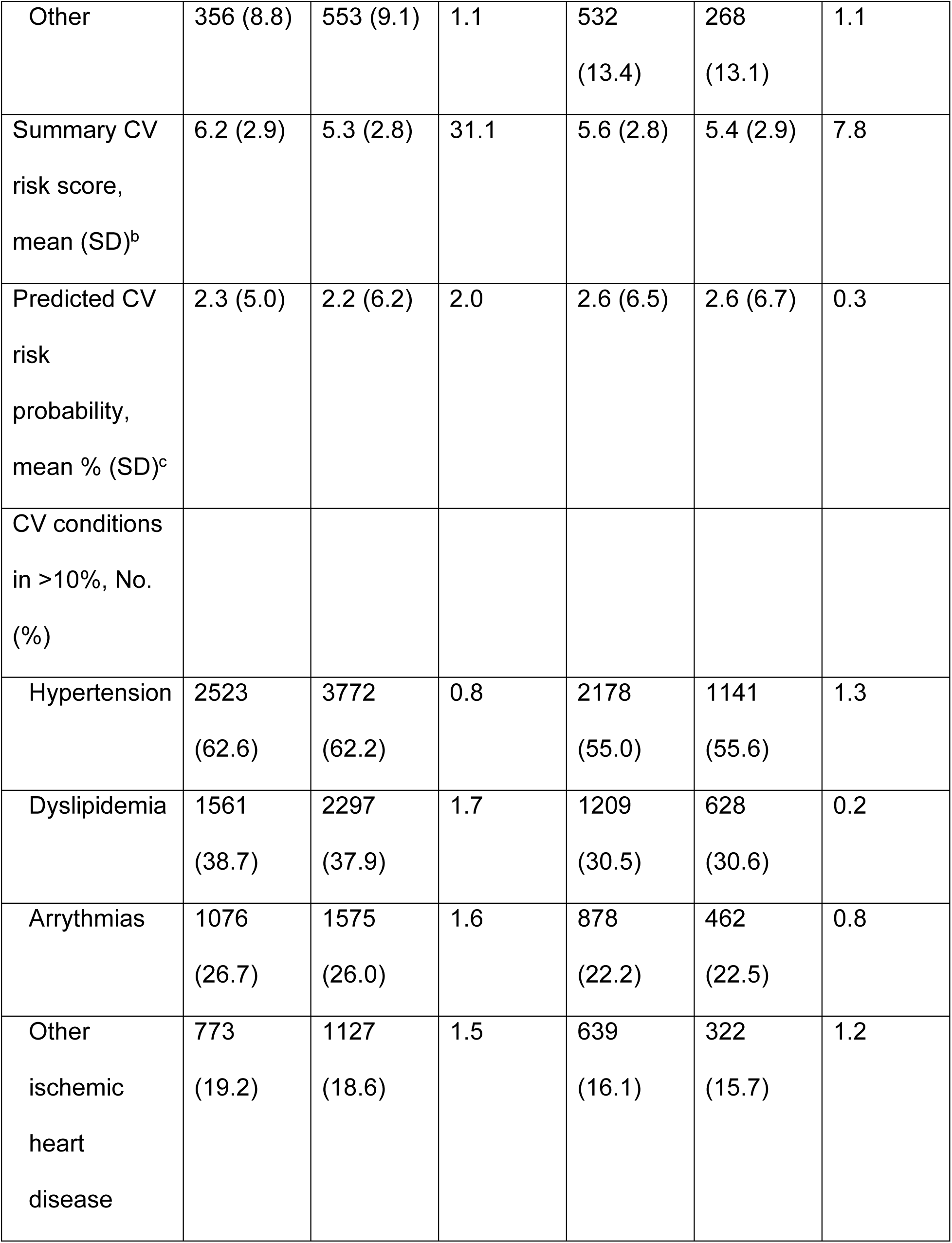
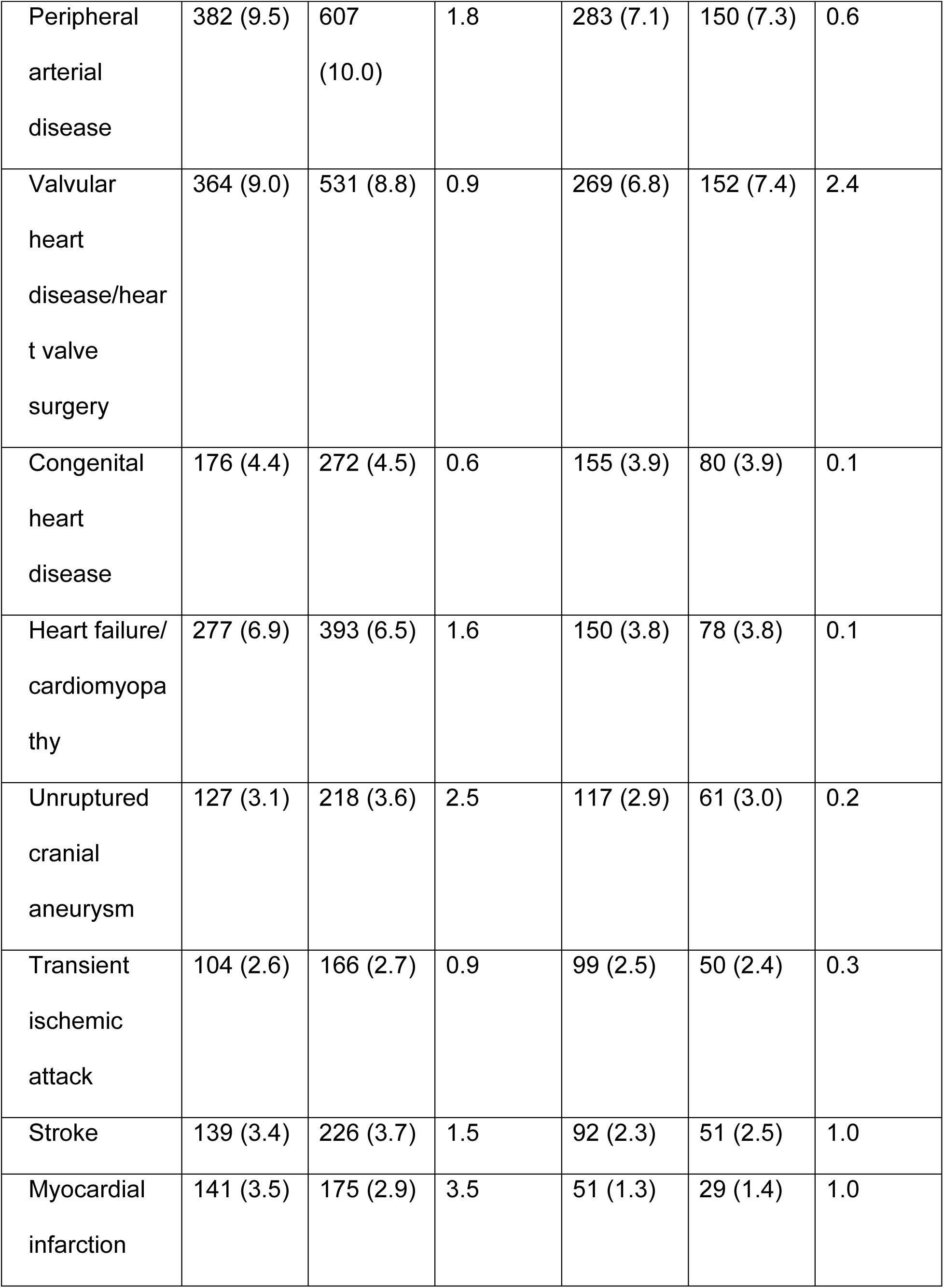
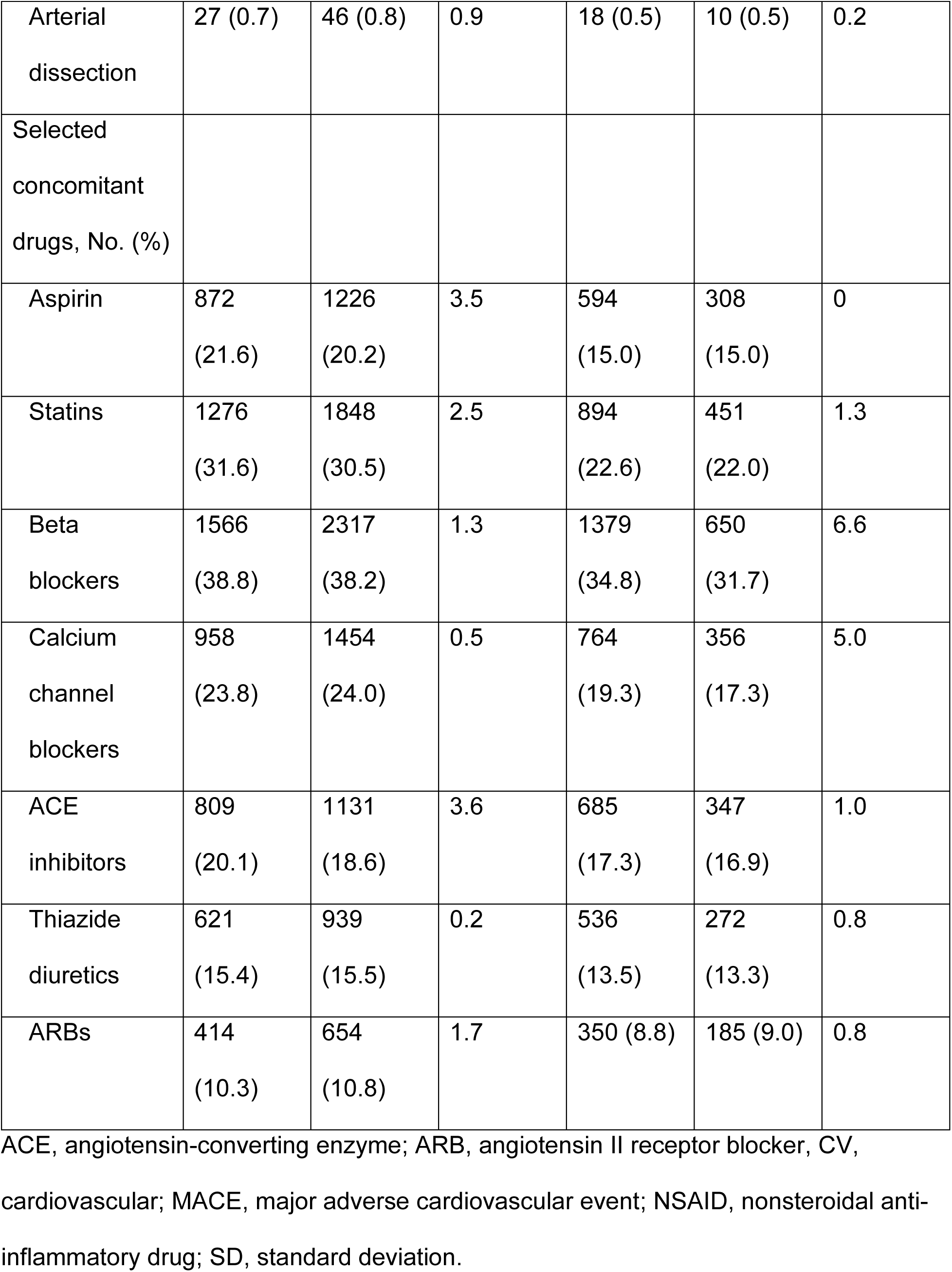
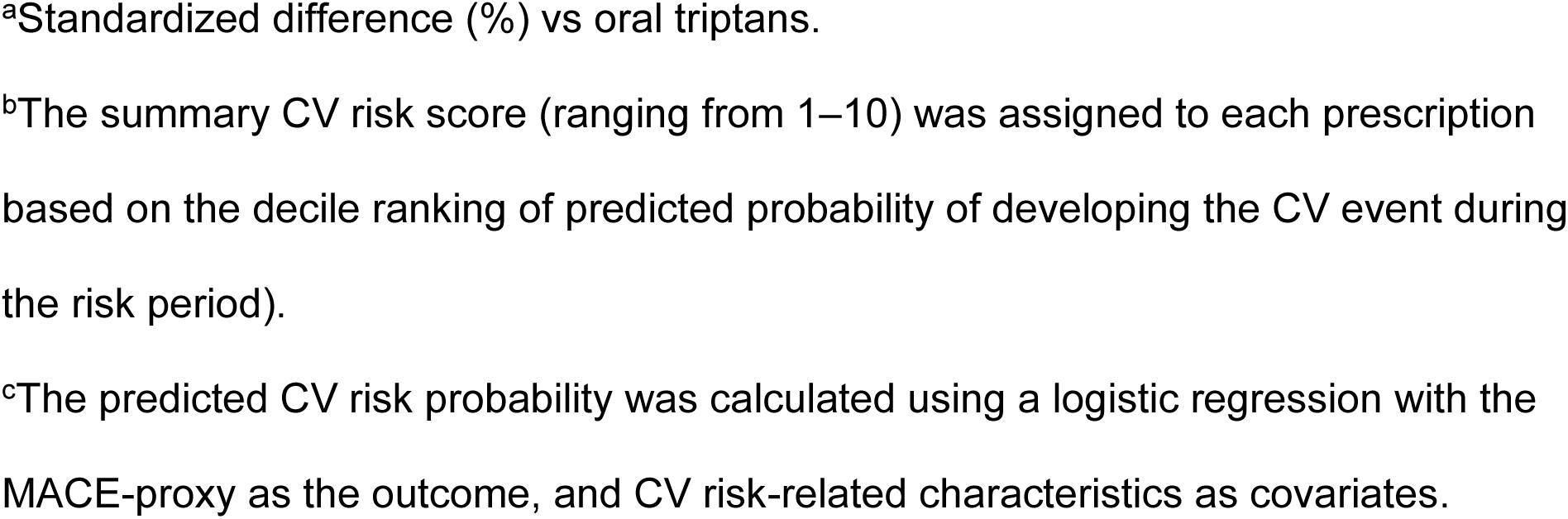
Population Characteristics (weighted sample)

|  | **Oral**<br>**triptans**<br>**(N=4033)** | **Opioids/**<br>**butalbital-based**<br>**therapies**<br>**(N=6066)** | **Standard**<br>**ized**<br>**differenc**<br>**e (%)<sup>a</sup>** | **Oral**<br>**triptans**<br>**(N=3962)** | **Non-**<br>**aspirin**<br>**NSAIDs**<br>**(N=2051)** | **Standard**<br>**ized**<br>**differenc**<br>**e (%)<sup>a</sup>** |
| --- | --- | --- | --- | --- | --- | --- |
| Mean age<br>(SD), years | 52.3<br>(14.7) | 52.3<br>(14.8) | 0.2 | 48.3<br>(14.2) | 48.5<br>(15.2) | 1.4 |
| Female, No.<br>(%) | 3538<br>(87.7) | 5270<br>(86.9) | 2.6 | 3443<br>(86.9) | 1781<br>(86.9) | 0.1 |
| Race/ethnicity,<br>No. (%) |  |  |  |  |  |  |
| White | 3092<br>(76.7) | 4631<br>(76.3) | 0.8 | 2705<br>(68.3) | 1399<br>(68.2) | 0.2 |
| Black/African<br>American | 300 (7.4) | 463 (7.6) | 0.8 | 358 (9.0) | 189 (9.2) | 0.6 |
| Hispanic/Lati<br>no | 153 (3.8) | 228 (3.8) | 0.2 | 208 (5.3) | 109 (5.3) | 0.2 |
| Asian | 61 (1.5) | 92 (1.5) | 0 | 70 (1.8) | 40 (1.9) | 1.2 |
| Unknown | 71 (1.8) | 98 (1.6) | 1.1 | 88 (2.2) | 46 (2.3) | 0.4 |
| Other | 356 (8.8) | 553 (9.1) | 1.1 | 532<br>(13.4) | 268<br>(13.1) | 1.1 |
| Summary CV<br>risk score,<br>mean (SD)<sup>b</sup> | 6.2 (2.9) | 5.3 (2.8) | 31.1 | 5.6 (2.8) | 5.4 (2.9) | 7.8 |
| Predicted CV<br>risk<br>probability,<br>mean % (SD)<sup>c</sup> | 2.3 (5.0) | 2.2 (6.2) | 2.0 | 2.6 (6.5) | 2.6 (6.7) | 0.3 |
| CV conditions<br>in >10%, No.<br>(%) |  |  |  |  |  |  |
| Hypertension | 2523<br>(62.6) | 3772<br>(62.2) | 0.8 | 2178<br>(55.0) | 1141<br>(55.6) | 1.3 |
| Dyslipidemia | 1561<br>(38.7) | 2297<br>(37.9) | 1.7 | 1209<br>(30.5) | 628<br>(30.6) | 0.2 |
| Arrhythmias | 1076<br>(26.7) | 1575<br>(26.0) | 1.6 | 878<br>(22.2) | 462<br>(22.5) | 0.8 |
| Other<br>ischemic<br>heart<br>disease | 773<br>(19.2) | 1127<br>(18.6) | 1.5 | 639<br>(16.1) | 322<br>(15.7) | 1.2 |
| Peripheral<br>arterial<br>disease | 382 (9.5) | 607<br>(10.0) | 1.8 | 283 (7.1) | 150 (7.3) | 0.6 |
| Valvular<br>heart<br>disease/heart<br>valve<br>surgery | 364 (9.0) | 531 (8.8) | 0.9 | 269 (6.8) | 152 (7.4) | 2.4 |
| Congenital<br>heart<br>disease | 176 (4.4) | 272 (4.5) | 0.6 | 155 (3.9) | 80 (3.9) | 0.1 |
| Heart failure/<br>cardiomyopathy | 277 (6.9) | 393 (6.5) | 1.6 | 150 (3.8) | 78 (3.8) | 0.1 |
| Unruptured<br>cranial<br>aneurysm | 127 (3.1) | 218 (3.6) | 2.5 | 117 (2.9) | 61 (3.0) | 0.2 |
| Transient<br>ischemic<br>attack | 104 (2.6) | 166 (2.7) | 0.9 | 99 (2.5) | 50 (2.4) | 0.3 |
| Stroke | 139 (3.4) | 226 (3.7) | 1.5 | 92 (2.3) | 51 (2.5) | 1.0 |
| Myocardial<br>infarction | 141 (3.5) | 175 (2.9) | 3.5 | 51 (1.3) | 29 (1.4) | 1.0 |
| Arterial<br>dissection | 27 (0.7) | 46 (0.8) | 0.9 | 18 (0.5) | 10 (0.5) | 0.2 |
| Selected<br>concomitant<br>drugs, No. (%) |  |  |  |  |  |  |
| Aspirin | 872<br>(21.6) | 1226<br>(20.2) | 3.5 | 594<br>(15.0) | 308<br>(15.0) | 0 |
| Statins | 1276<br>(31.6) | 1848<br>(30.5) | 2.5 | 894<br>(22.6) | 451<br>(22.0) | 1.3 |
| Beta<br>blockers | 1566<br>(38.8) | 2317<br>(38.2) | 1.3 | 1379<br>(34.8) | 650<br>(31.7) | 6.6 |
| Calcium<br>channel<br>blockers | 958<br>(23.8) | 1454<br>(24.0) | 0.5 | 764<br>(19.3) | 356<br>(17.3) | 5.0 |
| ACE<br>inhibitors | 809<br>(20.1) | 1131<br>(18.6) | 3.6 | 685<br>(17.3) | 347<br>(16.9) | 1.0 |
| Thiazide<br>diuretics | 621<br>(15.4) | 939<br>(15.5) | 0.2 | 536<br>(13.5) | 272<br>(13.3) | 0.8 |
| ARBs | 414<br>(10.3) | 654<br>(10.8) | 1.7 | 350 (8.8) | 185 (9.0) | 0.8 |

ACE, angiotensin-converting enzyme; ARB, angiotensin II receptor blocker, CV, cardiovascular; MACE, major adverse cardiovascular event; NSAID, nonsteroidal anti-inflammatory drug; SD, standard deviation.
<sup>a</sup>Standardized difference (%) vs oral triptans.
<sup>b</sup>The summary CV risk score (ranging from 1–10) was assigned to each prescription based on the decile ranking of predicted probability of developing the CV event during the risk period).
<sup>c</sup>The predicted CV risk probability was calculated using a logistic regression with the MACE-proxy as the outcome, and CV risk-related characteristics as covariates.

**Supplemental Figure 1.**
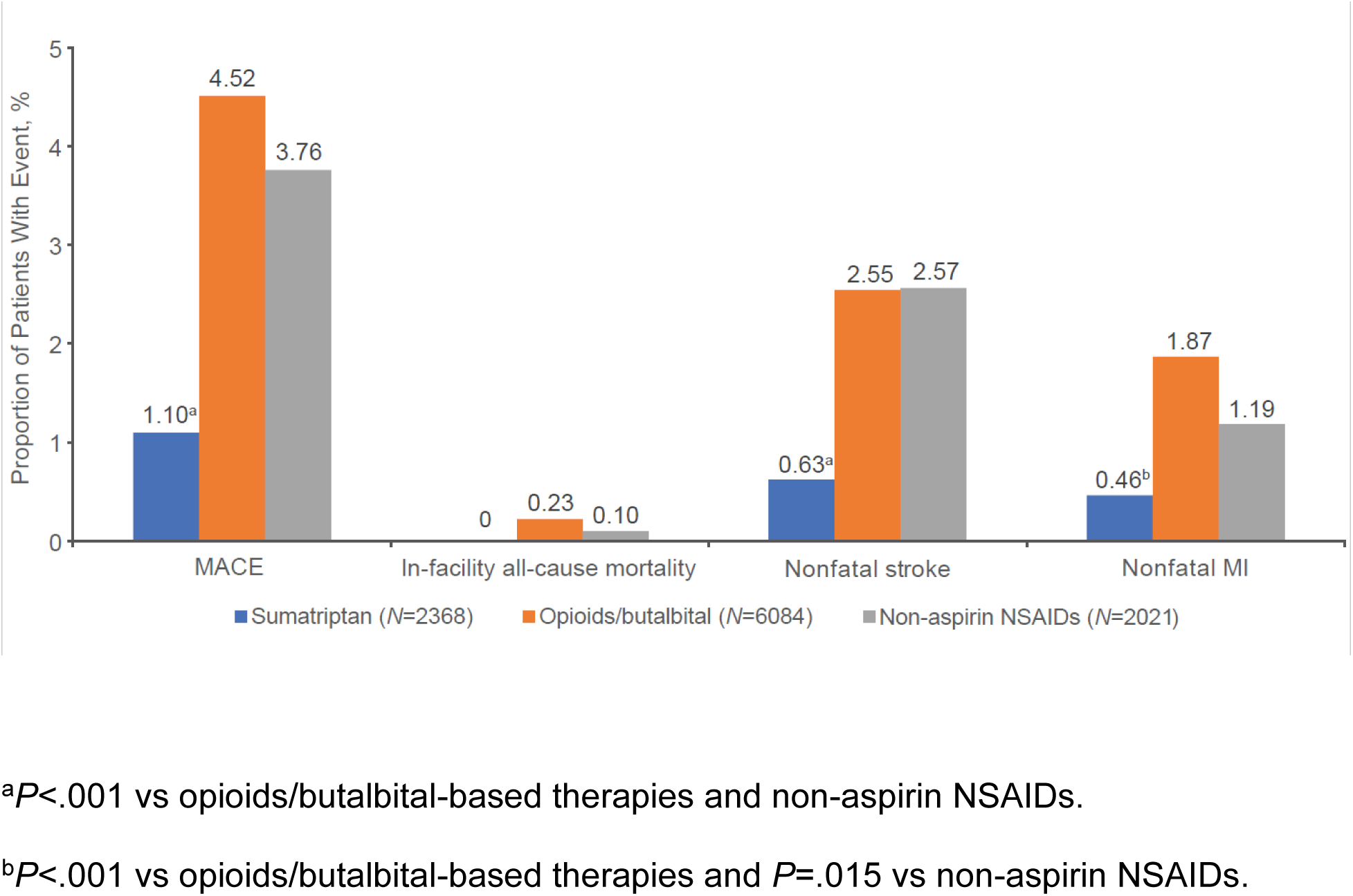
Summary of cardiovascular endpoints during the risk period (unweighted estimates): oral sumatriptan subgroup analysis. MACE, major adverse cardiovascular event (defined as first occurrence of any nonfatal MI, nonfatal stroke, or in-facility all-cause mortality); MI, myocardial infarction; NSAID, nonsteroidal anti-inflammatory drug.

**Supplemental Figure 2.**
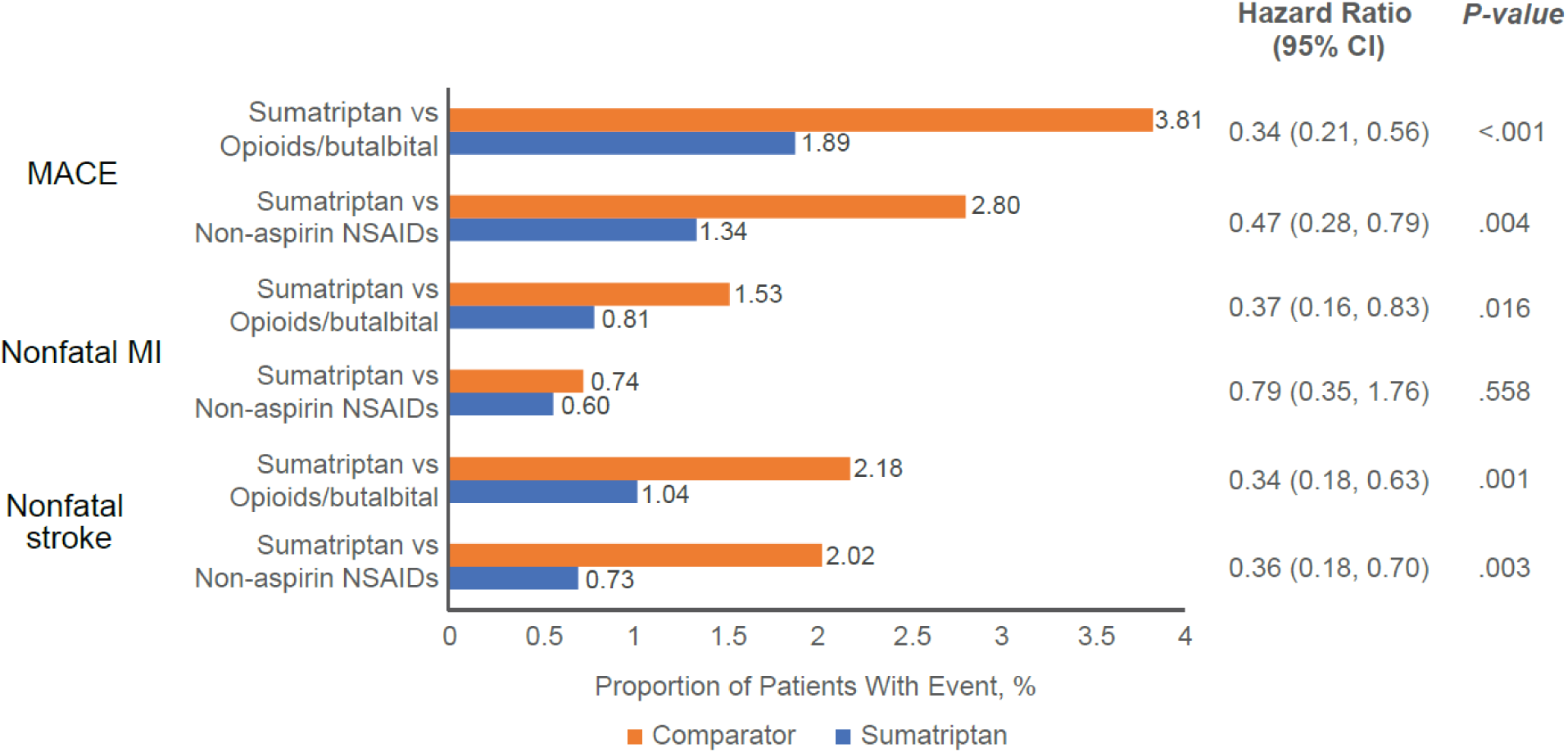
Proportion of patients with cardiovascular events during the risk period (weighted estimates): oral sumatriptan vs opioids/butalbital and non-aspirin NSAIDs (adjusted model). MACE, major adverse cardiovascular event (defined as first occurrence of any nonfatal MI, nonfatal stroke, or in-facility all-cause mortality); MI, myocardial infarction; NSAID, nonsteroidal anti-inflammatory drug.

